# Placental microRNAs associate with early childhood growth characteristics

**DOI:** 10.1101/2022.02.03.22270310

**Authors:** Elizabeth M. Kennedy, Karen Hermetz, Amber Burt, Dong Pei, Devin C Koestler, Ke Hao, Jia Chen, Diane Gilbert-Diamond, Usha Ramakrishnan, Margaret R. Karagas, Carmen J Marsit

## Abstract

Poor placental function is a common cause of intrauterine growth restriction, which in turn is associated with increased risks of perinatal morbidity, mortality and long-term adverse health outcomes. Our prior work suggests that birthweight and childhood obesity-associated genetic variants functionally impact placental function and that placental microRNA are associated with birthweight. To address the influence of the placenta beyond birth, we assessed the relationship between placental microRNAs and early childhood growth. Using the SITAR package, we generated two parameters that describe individual weight trajectories of children (0-5 years) in the New Hampshire Birth Cohort Study (NHBCS). Using negative binomial generalized linear models, we identified placental microRNAs that associate with growth parameters (FDR<0.05), while accounting for sex, gestational age at birth, and maternal parity. Genes targeted by the six growth trajectory-associated microRNAs are enriched (FDR<0.02) in growth factor signaling (TGF/beta: miR-1290; EGF/R: miR-155, Let-7c; FGF/R: miR-155; IGF/R: Let-7c, miR-155, miR-1290), cyclic AMP signaling (miR-1246), calmodulin signaling (miR-216a, miR-1246), and NOTCH signaling (miR-629). These pathways function in placental proliferation, differentiation and function. Our results support the hypothesis that fetal environment, specifically placental cellular dynamics and function guided by microRNA expression, can have impacts beyond birth, into early childhood.

## INTRODUCTION

The Developmental Origins of Health and Disease (DOHaD) posits that *in utero* exposures can induce permanent maladaptive changes that “program” the fetus to have increased risk of later-life diseases^22^. The placenta is the master regulator of the intrauterine environment and an ideal tissue in which to test the DOHaD paradigm. This ephemeral organ facilitates the exchange of gases, nutrients and waste for the developing embryo ^23,24^, in addition to aiding nutrient metabolism, and acting as an endocrine organ critical for early development. Thus, its functions are responsible for proper development and programming of the offspring.

MicroRNAs are 21-25 base pair non-coding RNAs that regulate gene expression post-transcriptionally via sequence complementarity to the 3’ untranslated region of mRNA transcripts. microRNA-mediated gene regulation is achieved through target mRNA translational inhibition or degradation^25^. MicroRNAs are proposed to regulate more than 50% of human genes ^25–28^, emphasizing their dynamic utility as post-transcriptional regulators of gene expression. Placental microRNAs shape placental development and function by targeting genes that regulate trophoblast proliferation and differentiation, apoptosis, invasion, cellular metabolism, as well as vasculo- and angio-genesis ^28^. In a study of placental microRNA sequencing data from more than 500 mother-infant pairs, in two independent cohorts, we have previously identified a group of placental microRNAs that are associated with birthweight. The most robust birthweight differentially expression microRNA, miR-532, was associated with the differential expression of placental adipokines, leptin and adiponectin receptor^1^.

The placental transcriptome may have lasting metabolic impact beyond birth. Using parallel genomic and transcriptomic data from the Rhode Island Child Health Study (RICHS), Peng et al. reports that DNA variants that predict placental gene expression level (eSNPs) for a given gene are over-represented among loci that associate with childhood obesity and BMI ^29^. Compared to eQTLs for seven adult tissues, placental eQTLs were most strongly enriched among results from GWASs of birthweight, childhood obesity, and childhood BMI. These results demonstrate that the placental transcriptional landscape can have a lasting impact on early childhood growth and metabolism^29^.

In this study, we pose the question: Do microRNAs that regulate the placental mRNAs have a lasting impact on childhood growth? To address this question, we model childhood weight from birth to five years of age in the New Hampshire Birth Cohort Study (NHBCS). Using parameters derived from individual growth curves, we relate placental microRNA expression, from small-RNA sequencing, with early childhood growth. We use *in silico* methods to predict mRNA targets of interesting microRNAs and pathway analysis to add biological context to our findings.

## MATERIALS AND METHODS

### Cohort

#### The New Hampshire Birth Cohort Study

NHBCS was initiated in 2009 and is an ongoing study comprised of a cohort of mother-infant pairs. Pregnant women between 18 and 45 years of age were recruited from the study’s participating prenatal care clinics in New Hampshire, USA. Women were included in the cohort if their primary source of drinking water was from an unregulated residential well, they had resided in the same household since their last menstrual period and had no plans to move before delivery. All participants provided written informed consent in accordance with the requirements of the Committee for the Protection of Human Subjects, the Institutional Review Board (IRB) of Dartmouth College. In this study, NHBCS participants were singleton pregnancies recruited between February 2012 and September 2013.

### Data collection

#### Anthropomorphic measures

NHBCS has parallel demographic and anthropomorphic measures for mothers and newborns, as well as placental microRNA transcript abundance. Up to 14 weight measurements between birth and five years of age were abstracted from pediatric medical records (Ages: birth, 2 weeks, 1 month, 2 months, 4 months, 6 months, 9 months, 12 months, 15 months, 18 months, 2 years, 3 year, 4 years and 5 years; Fig. 1). Study participants were excluded from the analysis if they had less than three separate weight observations, but were not excluded for intermediate missing observations (Fig. 1).

**Fig. 1.**
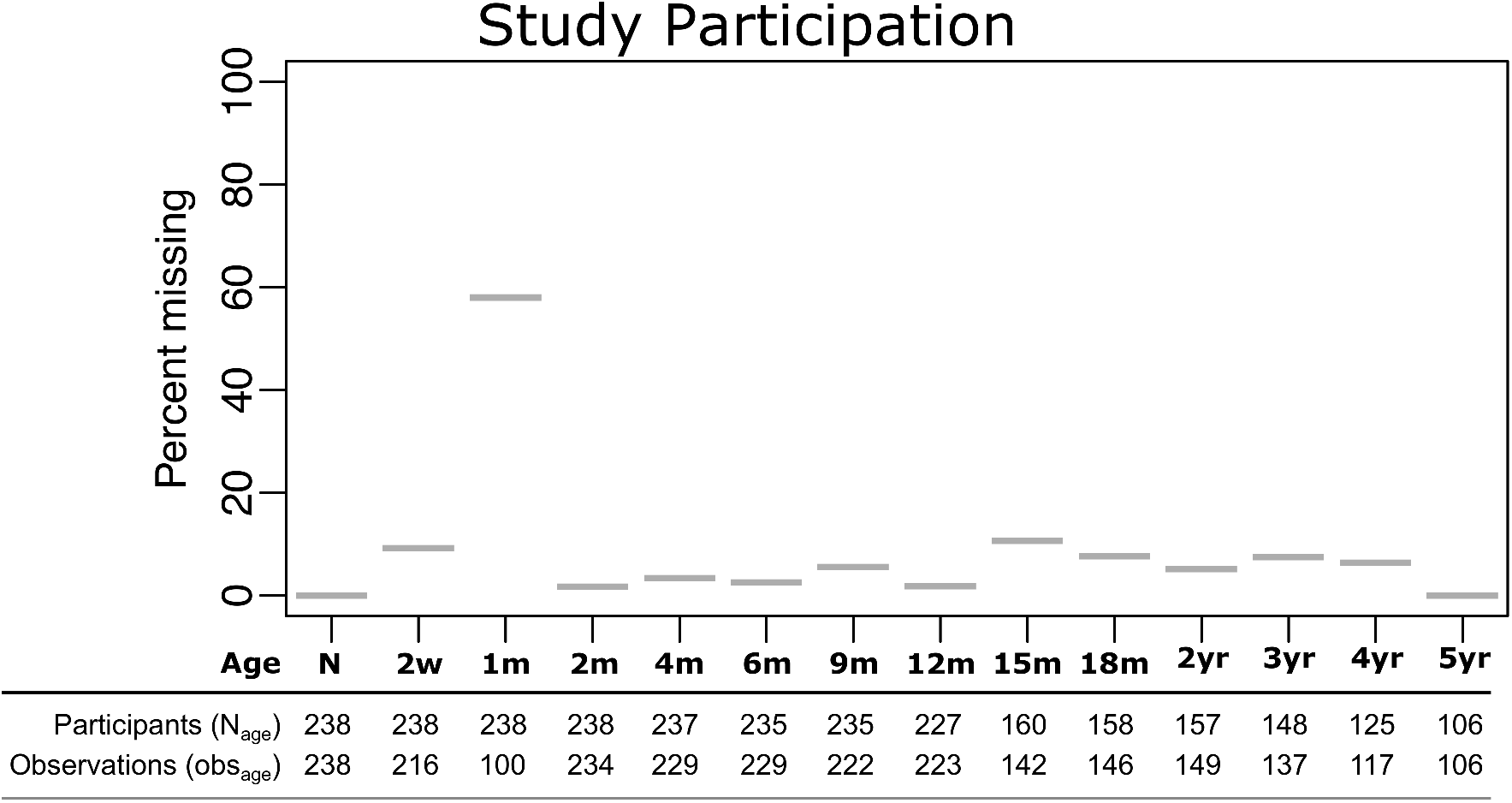
Participation in weight observation collection. Weights were collected at up to 14 separate time points for each child (x-axis). With the exception of the 1-month (1m) measure, missingness rate (y-axis) was low, meaning that observations at each measurement (obs_age_) remained high despite missing data after 12 months (12m, N_age_).

#### Additional study covariates

Gestational age and fetal sex were abstracted from newborn medical records. *z*-Scores were calculated for gestational age. Self-reported maternal parity was collected by questionnaire.

#### Tissue collection

Fetal placental samples were collected at delivery; sections were obtained two centimeters (cm) from the umbilical cord and free of maternal decidua. Collected tissue was immediately placed in RNA later solution (Life Technologies, Grand Island, NY, USA) and stored at 4 °C for at least 72 hours and stored at −80 °C.

#### microRNA isolation and sequencing

Total RNA was extracted from placenta using the Qiagen miRNeasy Mini Kit and a TissueLyser LT (Qiagen, Frederick, MD, USA) following manufacturer’s protocol. Briefly, 25-35 mg of placental tissue was placed in a 2 ml round bottom tube with 700 ul of Qiazol Lysing Reagent and one 5 mm stainless steel bead. The tissue was homogenized on the TissueLyser LT for 2 minutes at 40 Hz. The resulting homogenate was processed with the Qiagen miRNeasy Mini Kit and eluted in 30 µl RNase-free water. The RNA was quantitated on a NanoDrop 2000 (Thermo Fisher, Waltham, MA, USA) and quality checked on Agilent Bioanalyzer using the Agilent RNA 6000 Nano kit (Agilent, Santa Clara, CA, USA). Single end, 1 × 75 bp next generation sequencing of placental microRNA was performed by Qiagen Genomic Services (Frederick, Maryland).

#### smallRNA-Seq Processing and Quality Control

Raw FASTQ reads obtained from a total of 552 (RICHS 230, NHBCS 322) samples were subject to adaptor trimming with *cutadapt* v1.16 ^30^. The 3’ adaptor sequence (AACTGTAGGCACCATCAAT) was trimmed based on vendor’s recommendation (Qiagen). After adaptor trimming, *fastQC* v0.11.5 was used to process the trimmed reads and QC results were aggregated using *MultiQC* v1.5 for visualization ^31^. One sample failed QC and was removed. Then we used trimmed reads and *miRDeep2* to quantify microRNA ^32^. In short, *miRDeep2* was used to first perform alignment using *bowtie1* with human genome hg38 ^33^. The ‘Quantifier’ module in *miRDeep2* was used to obtain raw counts of microRNAs with *miRBase* version 22 ^34^.

#### Sample filtering and transcript filtering

Raw counts were imported into *DESeq2* for normalization and differential expression analysis. microRNAs with less than one count per million in more than 10 percent of samples were removed. Of the 2,656 microRNA transcripts that mapped, 777 remained after filtering, respectively.

#### Normalization

Filtered microRNA raw counts were imported to DESeq2 for normalization and differential expression analysis. For all data sets, parametric estimates of dispersion were calculated, and the median ratio method was used to estimate size factors for normalization for modeling with *DESeq2* ^35^. Normalized counts were exported from *DESeq2* for surrogate variable analysis. RICHS RNAseq count data are available via dataverse (https://doi.org/10.15139/S3/FUC5EW) and the methods for collection and processing are available elsewhere ^1^.

### Statistical analyses

#### Modeling childhood growth trajectory

The R package SITAR was used to model growth trajectory for all study participants with at least three weight observations (n = 238)^36^. Individual children’s growth curves were visually assessed for outlier measures. Three observations were removed (Fig. S1). The children’s weights were natural log transformed to meet the normality assumption of the SITAR method. Briefly, SITAR is a shape invariant model, with individual random effects for each child, that estimates an average growth curve across samples. A set of three parameters are generated for each individual that, through translation (vertical/horizontal shift) and rotation (counter/clockwise), transform the average growth curve to match each individual’s growth. The parameters, random effects, have mean zero and standard deviations estimated from the data. These parameters - *size, intensity* and *tempo*, are interpreted as percentiles relative to the average because the weights were natural log transformed. The *size* parameter represents average *size* for any child relative to the average child and is graphically represented as a vertical shift of the weight curve (Fig. 2). *Tempo*, the age at peak weight *intensity*, was not estimated for this analysis, since no measures were estimated around the peak of infant growth (6 weeks); *intensity* is expressed as a percentage deviation from mean intensity with higher values representing faster growth than average. Graphically, *intensity* represents a rotation of the weight curve (Fig. 2). We modeled the transformed weights using 10 degrees of freedom, and adjusted for child sex in the models that generated the *size* and *intensity* parameters.

**Fig. 2.**
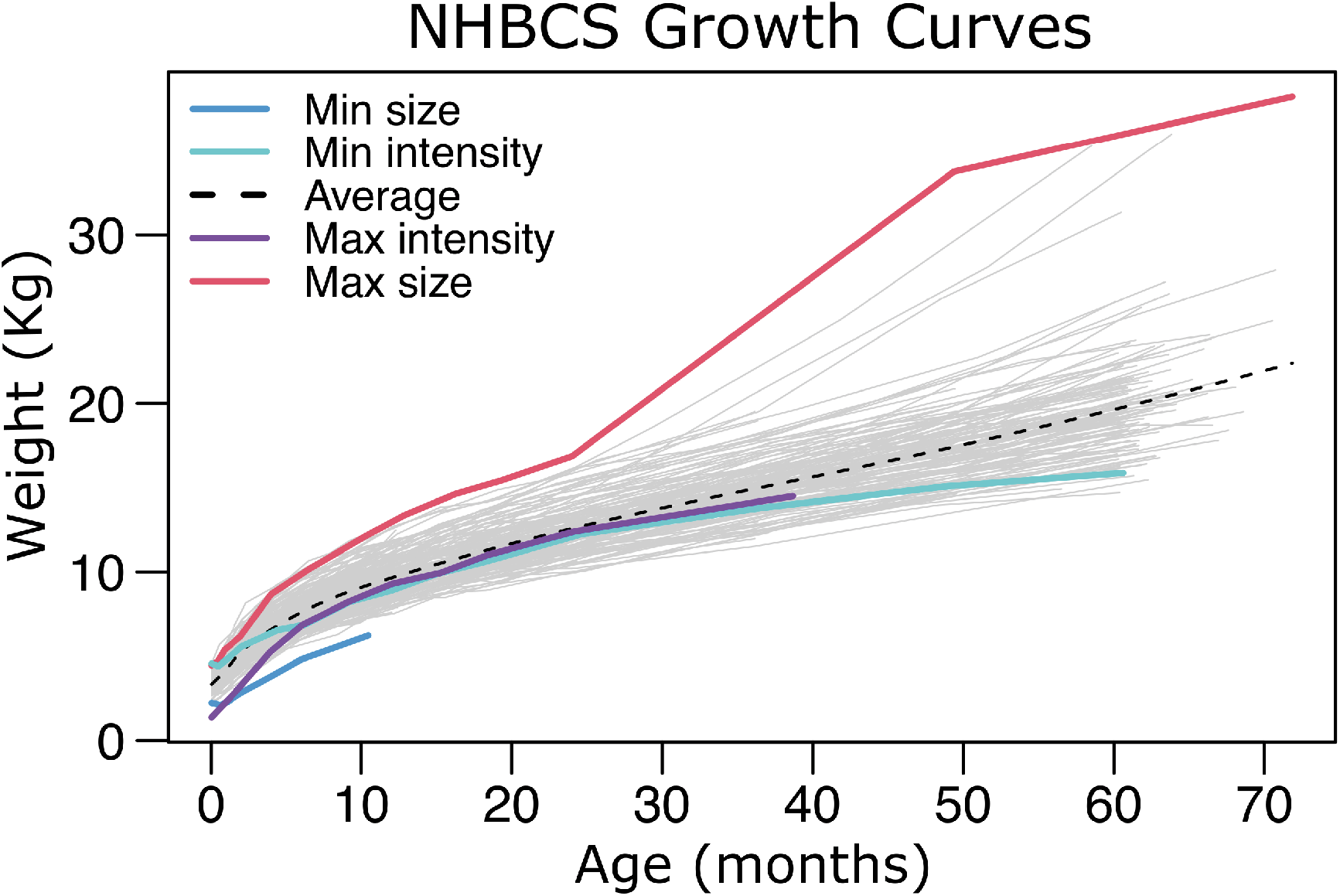
NHBCS Growth Curves. Unadjusted growth curves are plotted for all participants in this analysis (N = 238). The mean growth curve, calculated by SITAR, is plotted as a dashed line. The growth curves representing the minimum and maximum average *size* are plotted in blue and red, respectively. Deviations in average *size* appear as vertical shifts from the mean curve. The growth curves representing the minimum and maximum growth intensities are plotted in cyan and purple, respectively. Deviations in growth *intensity* appear as rotations (counter/clockwise) of the mean curve.

#### Surrogate variable analysis

To adjust for batch effects, cell-type heterogeneity and other unknown sources of technical variation, we estimated a surrogate variable from the normalized transcript reads via the *svaseq* function in the *sva* package that incorporates the Combat algorithm ^37,38^. In the *svaseq* function, the iteratively re-weighted least squares algorithm was used to estimate a surrogate variable based on empirically-derived control transcripts. The full model (mod argument) used for *svaseq* differed by application. The null model (mod0 argument) used all covariates except the outcomes variables - *intensity* and *size*.

#### Differential expression analyses

microRNA transcript counts were modeled using a negative binomial generalized linear model with significance testing for differentially expressed transcripts via Wald tests in *DESeq2* ^39^. The microRNA transcripts were regressed on either the *size* or *intensity* parameters from SITAR. The *intensity* analysis included covariates for parity and gestational age (z-score). Three samples were missing information for parity and were removed from the *intensity* analysis (n = 235). The *size* analysis included covariates for primigravida and maternal educational (beyond high school). Five samples were missing information for the maternal education variable and were removed from the *size* analysis (n = 230). One surrogate variable was also included as a covariate in both regression models (discussed in previous section). We considered microRNAs with a false discovery rate less than 5% to be differentially expressed with either *size* or growth *intensity*.

#### Sensitivity analysis

To assess potential confounding of DEmiR effects by pre-eclampsia, maternal pre-pregnancy BMI, gestational weight gain or birthweight, potential confounders were individually added to the DEA model in DESeq2. Effect estimates and standard errors were collected for *intensity* and *size* DEmiRs for comparison with original model statistics.

#### Target mRNA prediction

Potential mRNA targets of significant microRNAs were identified from mirDIP, an online database of human microRNA-target predictions ^40^. mirDIP integrates microRNA target prediction across 30 different resources, providing nearly 152 million human microRNA-target predictions. Using the individual source ranking and confidence measures, mirDIP assigns a unified rank and confidence score using the quadratic function. For this analysis, the predictions with the top 1% confidence scores (very high confidence) were returned for each microRNA^40^. The resulting microRNA-target mRNA pairs were carried forward in the analysis.

### Putative target filtering

#### Predicted

microRNA-target mRNA pairs were excluded if the mRNAs were not expressed in placenta in the RICHS whole transcriptome data.

#### Pathway analysis

Pathway over-representation analysis was conducted in consensuspathDB ^41^, which aggregates data from 12 separate pathway analysis databases. For each pathway gene set, consensus path DB calculates a p-value according to the hypergeometric test for the genes in both the putative target genes and the pathway gene set. All mRNAs not filtered for low reads in the RICHS whole transcriptome dataset were included as the background, or null distribution, for the test.

## RESULTS

This study analyzed data from 238 mother-infant pairs from the New Hampshire Birth Cohort (NHBCS). Parallel placental microRNA transcript abundance was available for this cohort, as well as up to 14 weight measures between birth and 5 years, collected from well-child-checks. Study participants were excluded from the analysis if they had less than three separate weight observations, but were not excluded for intermediate missing observations (Fig. 1). Observations as a fraction of participants in the study at each measurement remained high throughout the study, despite attenuated participation with time, especially after one year (Fig. 1). 48% of participants had at least 12 of the 14 potential observations and only 2% had less than 5 observations (median observations across participants is 11, ranged 3–14).

### Growth Trajectory modeling

We modeled childhood weight growth trajectory using SuperImposition by Translation And Rotation (SITAR)^36^. SITAR provided two parameters to describe each child’s growth. The *size* parameter is given as a percent, relative to the fitted weight curve for all participants, for the average *size* of each individual child. Higher *size* is graphically represented as an upward vertical shift in a child’s weight growth curve. The *size* parameter in this analysis ranged between -37% and 31% (Fig. 2, red and blue lines). The *intensity* parameter is expressed as a percentage deviation from the mean *intensity* for all participants, with higher values representing faster growth than average. Higher growth *intensity* is graphically represented as a counterclockwise rotation in a child’s weight growth curve. The *intensity* parameter for this analysis ranged between -9% and 12% (Fig. 1, purple and cyan lines).

The demographics of the participants for this study are shown in Table 1. Due to missingness in variables included in the models for the *intensity* and *size* differential gene expression analysis, three children were excluded in the *intensity* analysis (n = 235) and eight children were excluded in the *size* analysis (n = 230). The exclusion of children for the downstream analysis did not change the demographic distributions for each analysis (data not shown).

**Table 1.** Cohort demographics.

|  | Intensity<sup>a</sup> (n = 235) | Size<sup>a</sup> (n = 230) |
| --- | --- | --- |
| **MATERNAL CHARACTERISTICS** |  |  |
| Age - mean (range) | 31 (18 – 45) | 31 (18 – 45) |
| Education (>high school) - % (n) | 91 (206) | 90 (208) |
| Parity - mean (range) | 1 (0 – 4) | 1 (0 – 4) |
| Existing Diabetes | 0.4% (1) | 0.4% (1) |
| Gestational Diabetes | 5% (11) | 5% (12) |
| Pregnancy hypertension | 3% (7) | 3% (7) |
| Pre-eclampsia | 2% (4) | 2% (4) |
| Pre-pregnancy BMI - mean (range) | 26 (17 – 46) | 26 (17 – 46) |
| **Gestational weight gain category<sup>b</sup>** |  |  |
| Low<sup>c</sup> - % (n) | 7% (16) | 6.6% (15) |
| Normal<sup>d</sup> - % (n) | 26% (59) | 25.7% (58) |
| High<sup>e</sup> - % (n) | 67% (155) | 67.7% (153) |
| **INFANT CHARACTERISTICS** |  |  |
| Race (white) - % (n) | 100% (235) | 100% (235) |
| Gestational age (wks) - mean (range) | 40 (31 – 42) | 39 (31 – 42) |
| Birthweight (g) - mean (range) | 3464 (1380 – 4572) | 3468 (1380 – 4572) |
| **Birthweight group** |  |  |
| SGA<sup>f</sup> - % (n) | 4% (9) | 3% (8) |
| AGA<sup>g</sup> - % (n) | 86% (206) | 88% (202) |
| LGA<sup>h</sup> - % (n) | 10% (20) | 9% (20) |
| Sex (Female) - % (n) | 49% (116) | 49% (112) |

<sup>a</sup>Cohort characteristics and sample size for the SITAR *Intensity* and *Size* parameter analyses, respectively
<sup>b</sup>NHBCS statistics for gestational weight gain were based on the 230 samples with available measures in the *Intensity* parameter analysis and 226 for the *size* analysis.
<sup>c</sup>Low – Gestational weight gain below recommendation for pre-pregnancy BMI category<sup>42</sup>
<sup>d</sup>Normal – Gestational weight gain within recommendation for pre-pregnancy BMI category<sup>42</sup>
<sup>e</sup>High – Gestational weight gain above recommendation for pre-pregnancy BMI category<sup>42</sup>
<sup>f</sup>SGA – Infants born with a birthweight percentile $\leq 10^{43}$
<sup>g</sup>AGA – Infants born with a birthweight between the 10<sup>th</sup> and 90<sup>th</sup> percentiles<sup>43</sup>
<sup>h</sup>LGA – Infants born with a birthweight percentile $\geq 90^{43}$

### Differential microRNA expression analysis

To analyze the associations between growth trajectory parameters and placental microRNA expression, we performed differential expression analysis. For each growth trajectory parameter and microRNA transcript individually, transcript abundances were regressed on corresponding growth trajectory parameter using *DESeq2* ^39^.

Covariates were included in the model if they were associated with the variable of interest (*intensity* or *size*) in univariate regression analyses (Table 2). For the *intensity* analysis, models also included maternal parity, gestational age (z-scores) and one surrogate variable (to account for batch effects, cell type heterogeneity, and other sources of unknown variability). For the *size* analysis, primigravida, maternal educational attainment and one surrogate variable were included in the models. Of the 777 microRNA transcripts that passed quality control filtering for the analyses, five and one had an FDR < 0.05 for the *intensity* and *size* analyses, respectively (p-value < 1.4×10^−4^, Fig.s 3, S2). For the *intensity* and *size* analyses, respectively, two and one of those microRNAs had p-values less than the Bonferroni family-wise error rate threshold (p-value < 6.4×10^−5^, Fig.s 3, S2).

**Table 2.** Growth trajectory correlates.

| **Variable** | **Analysis** | **Estimate** | **Error** | **P-value** |
| --- | --- | --- | --- | --- |
| Pre-eclampsia | *Intensity* | 3.78 | 1.76 | 0.03 |
| Parity | *Intensity* | -0.64 | 0.22 | 4x10<sup>-3</sup> |
| Gestational weight gain category (low v normal) | *Intensity* | 2.10 | 0.99 | 0.03 |
| Gestational age | *Intensity* | -1.07 | 0.14 | 3x10<sup>-13</sup> |
| Birthweight percentile | *Intensity* | -0.06 | 0.01 | 1x10<sup>-11</sup> |
| Maternal education | *Size* | -0.45 | 0.22 | 0.04 |
| Primigravida | *Size* | 0.32 | 0.15 | 0.03 |
| Gestational weight gain category (high v normal) | *Size* | 0.4 | 0.15 | 9x10<sup>-3</sup> |
| Birthweight percentile | *Size* | 0.01 | 2x10<sup>-3</sup> | 7x10<sup>-8</sup> |

**Fig. 3.**
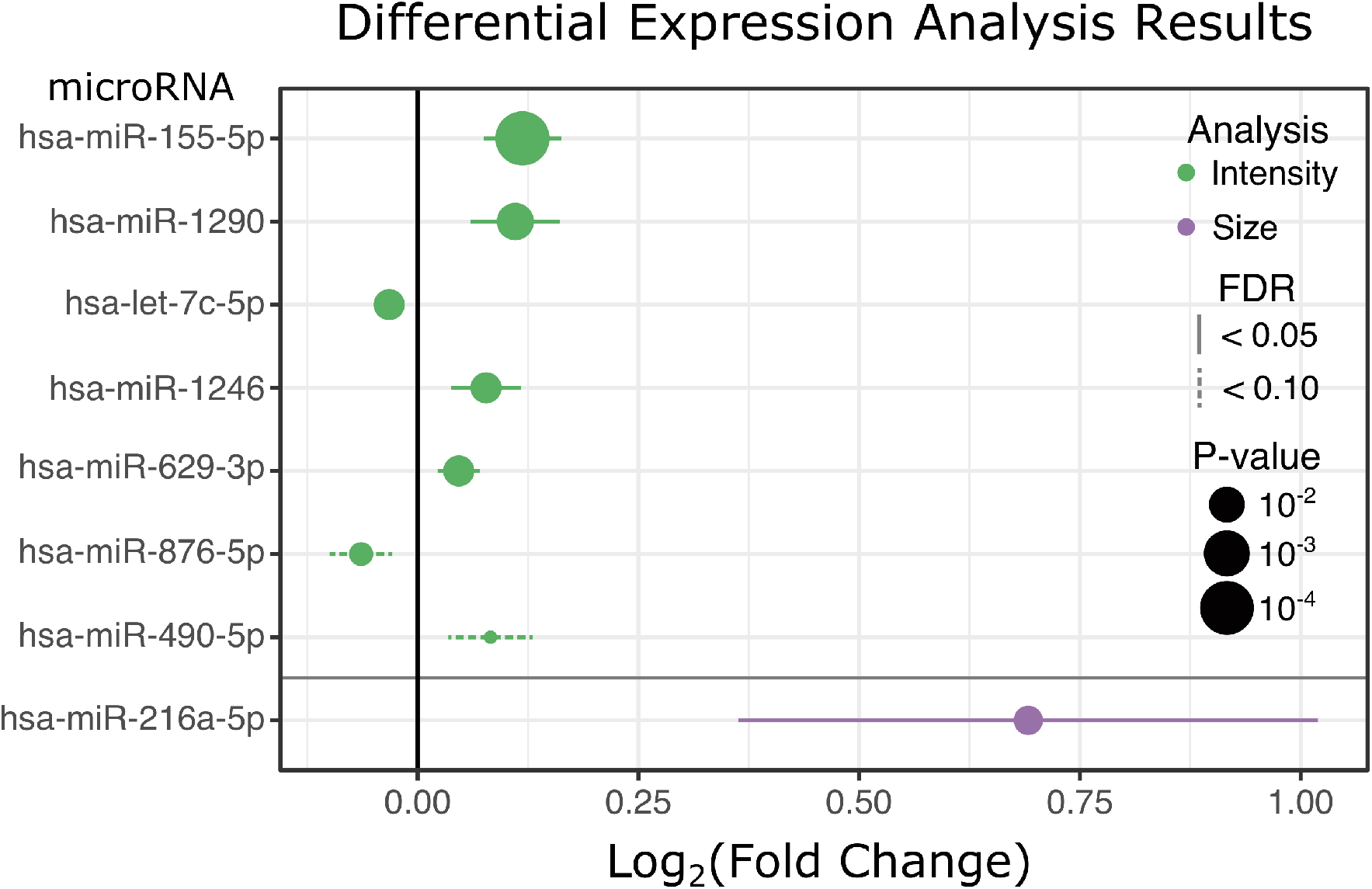
Differential expression analysis results. microRNAs with FDR<0.1 in either analysis are listed on the y-axis. Their log_2_ fold change for a one percent change in either growth *intensity* (green) or average *size* (purple) is on the x-axis. 95% confidence intervals are illustrated with either solid (estimate has FDR<0.05) or dashed (estimate has FDR<0.1) lines. Point size is proportional to -log10(P-value), such that larger points represent smaller p-values.

MicroRNAs that associate with *intensity* are hsa-miR-155-5p (estimate = 0.12; standard error: 0.02; FDR = 8.7×10^−5^), hsa-miR-1290 (estimate = 0.11; standard error: 0.03; FDR = 7.7×10^−3^), hsa-let-7c-5p (estimate = -0.03; standard error: 8×10^−3^; FDR = 0.02), hsa-miR-1246 (estimate = 0.08; standard error: 0.02; FDR = 0.02), and hsa-miR-629-3p (estimate = 0.05; standard error: 0.01; FDR = 0.02; Fig. 3). The *intensity* differentially expressed microRNAs (DEmiRs) have modest effects. One microRNA, has-miR-216a-5p, positively associates with average *size* (estimate = 0.69; standard error: 0.17; FDR = 0.03; Fig. 3). The estimates represent the log_2_ fold-change in microRNA abundance for one percent change in *intensity* or *size*.

Birthweight percentile was not included in the model because of its potential involvement in the causal path of growth trajectory. Of the microRNAs that were associated with growth trajectory with an FDR < 0.1, four (miR-155, miR-629, let-7c and miR-1246) were also associated with birthweight (FDR<0.1) in NHBCS ^1^. Because only four participating mothers had pre-eclampsia, it was not included in the primary analysis. However, the potential for pre-eclampsia and other potential confounders (pre-eclampsia, maternal pre-pregnancy BMI, gestational weight gain and birthweight) to attenuate significant associations (Fig. 3) between placental microRNAs and growth trajectory parameters was assessed in sensitivity analyses (Fig. S3). As expected for a variable along the causal path, the inclusion of birthweight as a covariate in the model strongly attenuated the estimated log_2_ fold change for *intensity* among microRNAs that are also associated with birthweight. Although miR-216a is not associated with birthweight, the addition of birthweight to the model associating transcript abundance to average *size* attenuated the observed association with *size*. Interestingly, the addition of categorical gestational weight gain (high, normal, low)^42^ as a covariate in the analysis of miR-216a-5p on average *size* resulted in a near doubling of the estimated log_2_ fold change of miR-216a-5p for each 1% change in average *size*. The inclusion of maternal pre-pregnancy BMI as a covariate decreased both the effect and the precision of the association of miR-216a-5p with average *size*.

### Growth trajectory DEmiR associated mRNAs

Bioinformatic targets of *Intensity-* and *size-* associated DEmiRs with an FDR < 0.05 were collected from the microRNA Data Integration Portal (mirDIP) ^40^. In order to enrich our targets with true microRNA-target pairs, we utilized total RNA abundance, assayed from 199 placentae using RNAseq in the RICHS cohort (https://doi.org/10.15139/S3/FUC5EW). mRNA transcripts were considered putative targets of trajectory DEmiRs if they were detectable in RICHS placenta samples. Using these criteria, we found 1,273 putative miR-155 targets, 313 miR-1290 targets, 1,256 let-7c targets, 138 miR-1246 targets, 38 miR-629 targets and 747 miR-216a targets. DEmiR mRNA targets were used in pathway analysis.

For each DEmiR, putative target genes were tested for pathway overrepresentation with consensuspathdb (CPDB), against all of the genes that passed QC in the RICHS whole transcriptome RNA-seq analysis ^41^. The most significant pathways are listed in **Table 3**. CPDB integrates pathways from 12 databases, meaning that similar results can be reported across sources. To further prioritize significantly overrepresented pathways, we generated term frequency matrices for all pathways for which microRNA targets made up more than 20% of pathway genes and for which the enrichment had an FDR less than 0.02. To assess the microRNA target genes leading the enrichments, a second term frequency matrix was made from the microRNA target genes present in the significant pathways. From these analyses, we found that FGFR1-4 and IGF/R signaling pathways were overrepresented among significant miR-155 pathways. The most frequently involved miR-155 target genes were PIK3CA and PIK3R1. For miR-1290, BMP and TGF/beta signaling were the most frequent pathways, with MAPK1 and SMAD2-4 the most frequent target genes. EGF and EGFR pathways were the most frequent among let-7a significant pathways, with MAPK1, PIK3CA and PIK3R1 as the most frequently involved gene targets. There was not repetition among the 13 significant miR-1246 pathways, though the most frequent genes were ADCY1 and PRKACB. Among mir-629 pathways, PTEN was the most frequent pathway, with TNRC6C and AGO1 and AGO4 were the most common gene targets. Finally for the only *size* DEmiR, miR-216a, regulation of MECP2 was most frequent among pathways, with miR-216a target CALM1-3 as the most frequent genes. When targets of positively-associated growth *intensity* DEmiRs (Fig. 3) were pooled for pathway analysis, we found that the most frequent pathway terms were FGFR1-4 and the most frequent gene targets were PIK3CA and PIK3R1.

**Table 3.** Top significant pathway for growth trajectory DEmiRs.

| **microRNA** | **Analysis** | **Pathway** | **q-value** | **% pathway**<br>**genes as targets** |
| --- | --- | --- | --- | --- |
| hsa-miR-155-5p | *Intensity* | FLT3 Signaling | 4x10<sup>-3</sup> | 42 |
| hsa-miR-1290 | *Intensity* | Regulation of cytoplasmic and<br>nuclear SMAD2/3 signaling | 2x10<sup>-4</sup> | 24 |
| hsa-let-7c-5p | *Intensity* | EGF-EGFR Signaling<br>Pathway | 2x10<sup>-5</sup> | 47 |
| hsa-miR-1246 | *Intensity* | L1CAM interactions | 7x10<sup>-3</sup> | 24 |
| hsa-629-3p | *Intensity* | Signaling by NOTCH | 6x10<sup>-3</sup> | 21 |
| hsa-miR-216a-5p | *Size* | EGFR1 | 3x10<sup>-3</sup> | 11 |

## DISCUSSION

In this study, we have described the microRNAs from human placenta that associate with growth trajectory from birth to five years old. We used a shape invariant model with random effects to generate two parameters that describe children’s growth *intensity* and average *size* during the observation period. We found evidence of microRNAs that vary with both growth parameters (DEmiRs). We narrowed bioinformatically predicted microRNA targets to only those in which the targeted mRNA was stably expressed in term placenta in the RICHS cohort. These putative DEmiR targets were used in pathway over-representation analysis.

Placental function, which underlies successful pregnancy and may have lasting influence, may be dictated by cellular dynamics – the balance of cell proliferation and differentiation among the terminal placental trophoblast lineages^44^. Recent evidence has suggested that upstream progenitor cells, with the ability to differentiate into cytotrophoblasts, syncytiotrophoblasts and extravillous trophoblasts are present in placenta to term^45^. Diminished abundance of these progenitor cells corresponds to placental dysfunction, like pre-eclampsia^45^.

The formation of the placenta relies on the maintenance and proliferation of human trophoblast stem cells, guided, in part by WNT and EGFR signaling^46^. In mice, this maintenance of stemness is reliant on the presence of FGF signaling^47^, however, this may or may not be the case in human placentae^46^. IGF is also important for survival and proliferation of trophoblast stem cells^44^. In our analysis, we see evidence that the EGFR (miR-155, let-7c), FGFR (miR155) and IGFR (let-7c, miR-155, miR-1290) signaling pathways are influenced by growth trajectory microRNAs (Fig. 4).

**Fig. 4.**
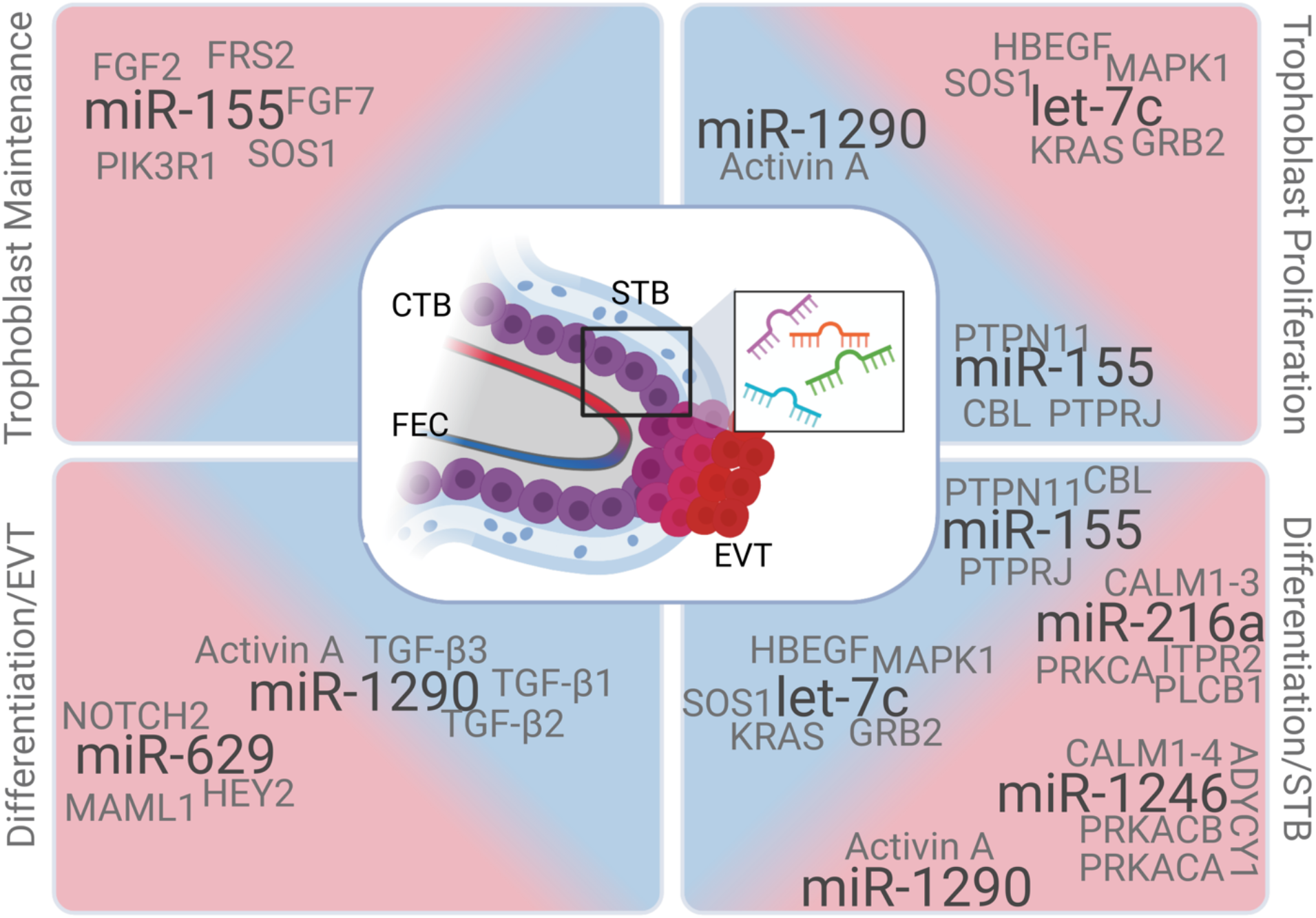
The potential roles of growth trajectory microRNAs in the cellular dynamics of placental trophoblasts. A terminal placental villous is illustrated in the center of the Fig., representing the involved placental cell types from which microRNAs in our study could arise: fetal endothelial cells (FEC), cytotrophoblasts (CTB), syncytiotrophoblasts (STB) and extravillous trophoblasts (EVT).Growth trajectory microRNAs are listed (in black) along with their influential targets (gray) in the processes we predict they may influence (trophoblast stem maintenance, proliferation and differentiation to the syncytiotrophoblast (STB) or extravillous trophoblast (EVT) terminal lineage). microRNAs and their putative targets in the central blue or outer red areas are predicted to encourage or inhibit the given process, respectively. Created with BioRender.com.

Multipotent villous cytotrophoblasts fuse to form the epithelial, multinucleated syncytiotrophoblast, which facilitates exchange of gases, nutrients and waste for the growing embryo, as well as acting as an endocrine organ to balance the needs of the mother and fetus^48^. Syncytialization is mediated by cAMP and rising intracellular Ca+, as well as continued EGF signaling and Activin A^46,49^. In this analysis, growth trajectory microRNAs are predicted to target components of the cAMP (miR-1246), calmodulin (miR-216a, miR-1246), EGFR (miR-155, let-7c) pathways, as well as Activin A (miR-1290; Fig. 4).

Cytotrophoblasts undergo a process similar to the epithelial-to-mesenchymal transition to form the invasive extravillous trophoblast that connects the placenta with the maternal decidua and myometrium to establish maternal blood supply^48^. Signaling through NOTCH1/2 and TGFβ family members (BMP2, Activins) mediate differentiation to the invasive extravillous cytotrophoblast^50–52^. The role of the TFG-beta superfamily in trophoblast differentiation is complicated, as many of the pathway mechanisms overlap^53^, yet TGF-beta1/2/3^53^ and NODAL^54^ inhibit the invasive phenotype and favor syncytialization while BMP2 and Activin A/B/AB all favor the invasive phenotype^50,51^. In this analysis, we found evidence that growth trajectory associated microRNAs target components of the NOTCH1/2 (miR-629) and TGFβ superfamily (miR-1290) signaling cascades (Fig. 4).

Although these processes described above were primarily described in first trimester placentae and cell culture, the placental cytotrophoblasts continue to maintain the syncytiotrophoblast and extravillous trophoblasts to term^45^. The discovery of trophoblast progenitor cells in term placenta suggests that some of the signaling pathways relevant in early gestation, remain important later in gestation. Within this assumption, our findings suggest that microRNAs influence the cellular dynamics of placenta at term, which in turn regulate the plasticity and efficiency of the placenta to support fetal growth. This initial growth primes the neonate’s metabolism for early growth patterns. However, it is also likely that some of the growth factors under microRNA influence have specific functions at term that are in addition to or exclusive of their functions in early gestation. For instance, while EGF signaling guides trophoblast maintenance, proliferation and differentiation in early gestation, it may stimulate the release of hormones to the maternal and fetal circulations at term^44^.

Some of the growth trajectory-associated microRNAs described in this analysis have been previously associated with birthweight in the NHBCS^1^ including miR-155-5p, mir-629-3p, let-7c and miR-1246. Birthweight was not included in our models, because it is an upstream measure of weight, and included as the initial weight in the growth trajectory analysis. Interestingly, the association of miR-1290 with growth trajectory is not sensitive to the inclusion of birthweight to the model, suggesting that placental miR-1290 relates to early childhood growth independently of birthweight.

Growth trajectory-associated microRNAs have also been associated with placental characteristics in other studies. In cultured extravillous trophoblasts, miR-155 inhibits cell proliferation by down-regulating cyclin D1/p27 ^55^. In cultured cytotrophoblasts, Let-7c is associated with reduced proliferation potential and syncytialization ^56,57^. Let-7c is associated with the WNT/β-catenin signaling pathway in other progenitor cell types^58^. In cultured extravillous trophoblasts, miR-1290 promotes rearrangement of maternal endometrium via placental exosomes ^59^. miR-1246 has been associated with syncytialization and targeting inhibitors of the WNT/β-catenin signaling pathway^60^.

To our knowledge, this is the first study to examine the relationships between human placental microRNAs and early childhood growth trajectory. Our results suggest that placental microRNAs effect signaling cascades central to trophoblast proliferation, differentiation and function. Most importantly, our results underscore the importance of placental function and the intrauterine environment in establishing early growth trends.

Our findings should be interpreted within the context of this study’s limitations. This is an observational study in which RNA was assayed from term placentae. Thus, we cannot conclude that our results are representative of microRNA associations throughout development. We used DESeq2 to identify associations between miRNA expression and childhood growth, where childhood growth was determined using child-specific estimates of *size* and *intensity* derived using SITAR. In DESeq2, microRNA abundances (dependent variable) are regressed on *size* or *intensity* (independent variable), allowing us to examine the association of early childhood growth on placental microRNA expression. Although the models employed in DESeq2 are temporally reversed in terms of what is being modeled as dependent and independent variables, DESeq2 represents the best choice for modeling gene expression data as it accounts for the over-dispersion of RNA-sequencing count data. While the estimates generated by these models may be less intuitive and temporally reversed from our hypothesis, the associations between placental microRNA expression with growth trajectory patterns ascertained using such models are nevertheless valid. We adjusted for likely confounders in our study, but cannot rule out the possibility that unmeasured or residual confounding remains in our analysis. Potential confounders are maternal BMI, gestational weight gain and pre-eclampsia. Sensitivity analysis suggests that the addition of these variables to our models would have little to no effect on our conclusions, though some of our top findings are sensitive to one or both. To limit unknown statistical confounding (e.g. differing cellular composition between individuals or population stratification), our models are adjusted using surrogate variable analysis. This is a data-driven approach that may incorrectly estimate confounding elements, adding variability to our model. Recent research indicates that SVA is one of the more robust and reliable methods in studies such as ours ^61^. Lastly, the cohort utilized in this study consisted predominantly of healthy white mothers from a rural New England region of the United States, potentially limiting the generalizability of our findings.

## Data Availability

All data produced in the present study are available online at https://doi.org/10.15139/S3/FUC5EW or upon reasonable request to the authors

## AUTHOR CONTRIBUTIONS

EMK, DCK, KH, JC, DG-D, MRK and CJM conceptualized and designed the study. KH, AB and DP acquired data. EMK analyzed and interpreted the data. EMK Drafted the article. KH, AB, DP, DCK, KH, JC, DG-D, UR, MRK and CJM critically reviewed and carefully revised the article. All authors approved of the version to be published.

## ETHICAL STANDARDS

All participants provided written, informed consent and all protocols were approved by the IRBs at the Women & Infants Hospital of Rhode Island, Dartmouth College and Emory University, respectively.

