## Supplemental Figures for "Placental microRNAs associate with early childhood growth characteristics"

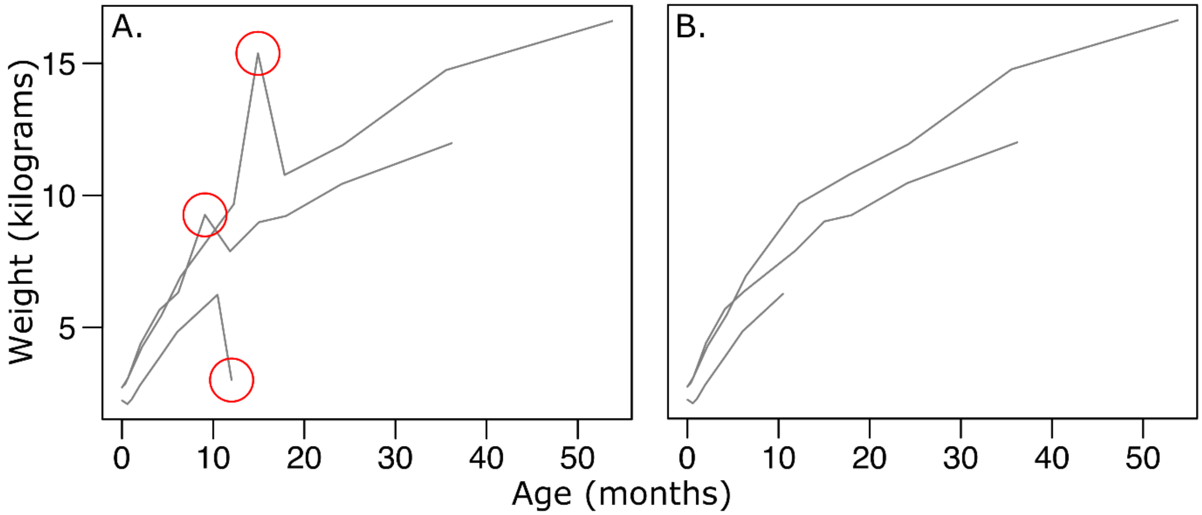


**Figure S1. Outlier weights in the NHBCS growth data.** Outlier weights (panel A; circled in red) were visually identified and removed from the data (panel B shows curves after outlier measure removal).

a.

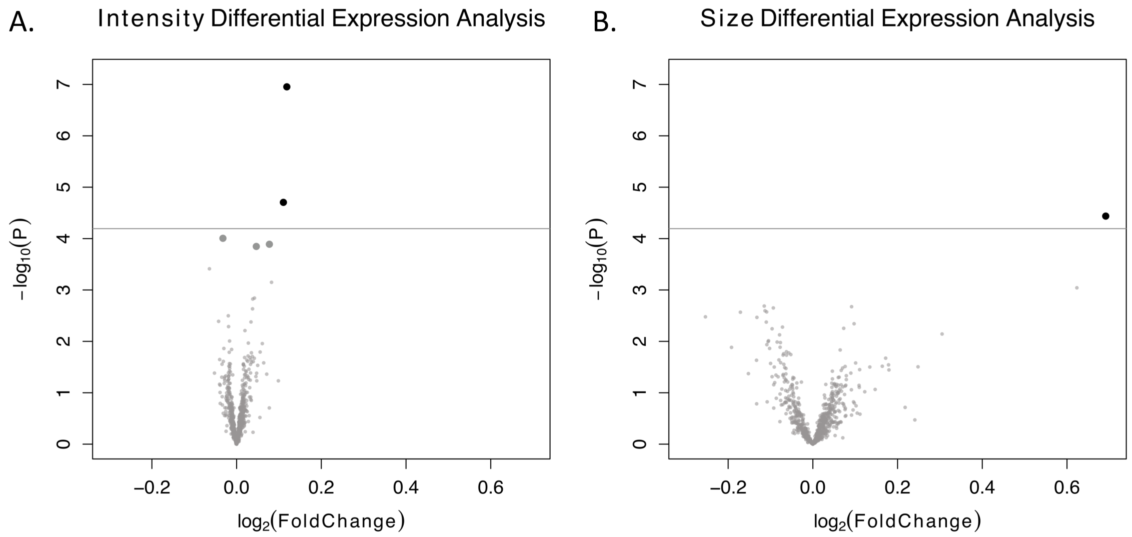


**Figure S2. Placental *microRNA association with growth trajectory parameters: intensity and size*.** Volcano plots illustrate the results of the differential expression analyses. On the y and x axes, -$\log_{10}$(p-values) in the association of each microRNA with *intensity* (A) or *size* (B) and effect estimates, or the $\log_2$ fold change in each microRNA per one percent change in *intensity* (a) or *size* (b), are shown respectively. The horizontal lines represent the Bonferroni threshold ($\log_{10}$(0.05/M), where M is the number of microRNAs tested). (A) 5 microRNAs are significantly (FDR < 0.05) associated with *intensity* (large gray points), with two microRNAs significant after Bonferroni correction (black points). (B) 1 microRNA is significantly associated with *size* after Bonferroni correction.

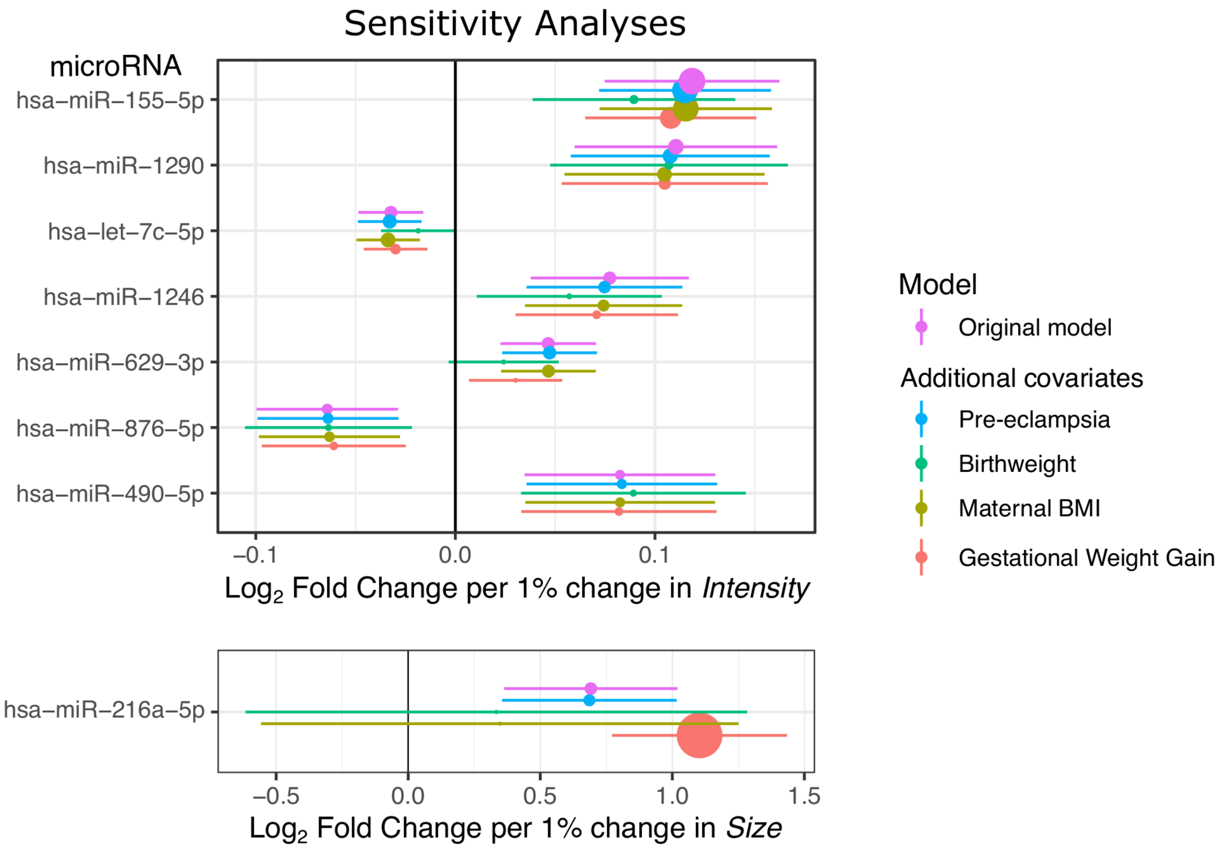


**Figure S3. Sensitivity analyses.** For each microRNA with FDR < 0.1 in either the growth *Intensity* or average *Size* analyses (y-axis), the estimates (x-axis) and 95% confidence intervals are given for the original model (pink) as well as with the indicated addition of either eclampsia (blue), birthweight (green), maternal pre-pregnancy BMI (brown) or gestational weight gain category (orange). Dot size is proportional to the -$\log_{10}$(p-value), such that larger dots represent more significant estimates.
